# Changing patterns of nicotine product use and nicotine dependence among US high school students: the National Youth Tobacco Survey, 2014-2023

**DOI:** 10.1101/2024.11.06.24316813

**Authors:** Sarah E. Jackson, Jamie Brown, Harry Tattan-Birch, Martin J. Jarvis

**Affiliations:** Department of Behavioural Science and Health, University College London, London, UK; SPECTRUM Consortium, UK

**Author notes:** Corresponding author: Dr Sarah Jackson, Department of Behavioural Science and Health, University College London, 1-19 Torrington Place, London WC1E 7HB, UK.

**Keywords:** dependence, smoking, e-cigarettes, tobacco products, adolescents, nicotine

## Abstract

**Background:** Concerns have been raised that e-cigarettes have created a new generation of people addicted to nicotine. This study aimed to examine changes in the proportion of US high-school students reporting symptoms of nicotine dependence over the past decade, in the context of changing patterns of nicotine product use.

**Methods:** Repeat cross-sectional analyses of a nationally-representative sample of 107,968 US high-school students (14-18y) participating in the 2014-2023 National Youth Tobacco Surveys. Nicotine product use was categorised based on self-reported past-30-day use of cigarettes, other combustible tobacco, smokeless/non-combustible products, and e-cigarettes. Nicotine dependence was operationalised as (i) strong past-30-day cravings to use tobacco and (ii) wanting to use nicotine products within 30 minutes of waking.

**Results:** Past-30-day use of any nicotine product decreased from 24.5% [22.5-26.6%] to 19.6% [16.8-22.4%] between 2014 and 2017, increased sharply reaching 31.4% [29.0-33.7%] in 2019, then fell to the lowest level at 12.5% [10.9-14.1%] by 2023. The proportion who reported symptoms of nicotine dependence was substantially lower, but followed a similar pattern of changes over time. For example, the proportion reporting strong cravings decreased from 7.8% [6.6-9.0%] to 5.5% [4.3-6.7%] between 2014 and 2017, increased to 7.9% [6.8-9.0%] between 2017 and 2018 and remained stable up to 2020, then fell to the lowest level at 2.5% [1.9-3.1%] by 2023. Use of cigarettes fell considerably across the period (from 9.0% [7.9-10.3%] to 1.8% [1.4-2.4%]); this was the product category consistently associated with the highest levels of dependence. The proportion using only e-cigarettes increased rapidly between 2017 and 2019 (from 5.4% [4.2-6.8%] to 17.0% [15.3-18.7%]) then fell to 6.7% [5.6-7.9%] by 2023; symptoms of nicotine dependence within this group increased non-linearly over time with increases through to 2022 before possible declines in 2023.

**Conclusions:** The sharp rise in the prevalence of nicotine product use (in particular, e-cigarettes) among US high-school students in the late 2010s was short-lived and was not accompanied by a sustained increase in the overall population burden of nicotine dependence. By 2023, both nicotine product use and nicotine dependence had reached historic lows.

## Introduction

Youth use of tobacco products is a long-standing public health concern.^1^ Rates of smoking have decreased steadily since the late 1990s among young people in the United States (US).^2,3^ However, a substantial rise in e-cigarette use (‘vaping’) among young people since 2017 has seen the overall decline in use of nicotine products reverse.^4^ This has caused concern that e-cigarettes are undoing decades of progress and creating a new generation of people addicted to nicotine.^5^ It is important to understand how far this is supported by evidence, to inform policy decisions around the regulation of e-cigarettes and other nicotine products. In this study, we examine how levels of nicotine dependence are changing among high-school students in the US in the context of evolving patterns of tobacco and nicotine product use.

In the US, youth vaping prevalence increased rapidly between 2017 and 2019. According to the 2019 National Youth Tobacco Survey (NYTS; an annual, cross-sectional, school-based survey), 31.2% of high-school students (grades 9-12; ages 14-18) had used an e-cigarette in the past 30 days,^6^ up from 11.7% in 2017.^7^ As of 2019, the NYTS data showed no corresponding large rise in the proportion of high-school students reporting symptoms of nicotine dependence.^8^ However, some researchers have raised concerns that e-cigarette users may not have interpreted questions about dependence on tobacco products as applying to them (despite this being explicitly stated in the survey),^9^ which warrants further investigation.

Since 2019, the vaping landscape has changed. There was an outbreak of acute lung injuries among (predominantly young) people using vaping products in 2019,^10^ which was initially attributed to nicotine-containing e-cigarette use before the cause was identified as inhalation of vitamin E acetate, found in illicit cannabis vaping products but not in nicotine e-cigarettes.^11^ Following the outbreak, young people were exposed to increased negative news stories about vaping and their harm perceptions of e-cigarettes worsened.^12^ There have also been regulatory changes in the US, including bans on vaping products sold by Juul, the brand associated with the rapid rise in vaping among young people; state- and local-level bans on flavours and/or online sales; and in some places an outright ban on the sale of all vaping products.^13–15^

Perhaps as a result of these factors, vaping prevalence among young people has fallen substantially over the past few years.^4^ The most recent NYTS data show prevalence of past-30-day e-cigarette use among US high-school students fell by almost two-thirds between 2019 and 2023, from 27.5%^6^ to 10.0%.^16^ There was a similar decline in the proportion reporting past-30-day use of any nicotine product, from 31.2%^6^ to 12.6%.^16^

However, newer vaping products that use salt-based nicotine e-liquids deliver nicotine more efficiently than older-generation devices,^17,18^ potentially leading to increased dependence among the remaining pool of vapers. Recent evidence from the International Tobacco Control (ITC) Youth Tobacco and Vaping Study show markers of dependence increased between 2017 and 2022 among 16-19-year-olds in the US, Canada, and England who vape, reaching levels comparable to cigarette dependence among those who smoke.^9^ This raises questions as to what has happened to the population burden of nicotine dependence among young people, in the context of decreased prevalence of vaping but potentially increased dependence among those who vape.

This study used NYTS data collected between 2014 and 2023 to describe changes in nicotine product use among high-school students and to examine the extent to which symptoms of nicotine dependence changed during this period, overall and within users of different nicotine product categories (e-cigarettes only, smokeless but no combustibles, combustibles but no cigarettes, and cigarettes). We also provided estimates accounting for potential underreporting of symptoms of nicotine dependence by students using e-cigarettes only.

## Methods

### Design

We used cross-sectional data from the NYTS collected annually between 2014 and 2023. Details of the NYTS methodology have been published in full elsewhere.^19^ Briefly, a three-stage cluster sampling procedure is used to recruit a nationally representative sample of students in grades 6–12. Participants provide data on a range of variables relevant to tobacco use via an anonymous questionnaire (administered in paper-and-pencil form up to 2018 and online from 2019 onwards).

We analysed data from high-school students (grades 9–12; age 14-18) surveyed between 2014 and 2023, because questions on nicotine dependence were consistently included in each year during this period. The total sample size was 107,968 (mean [SD] number of participants per year = 10,797 [2,179]).

### Measures

#### Nicotine product use

Nicotine product use was defined as any use of the following four categories on at least one of the past 30 days: cigarettes, other combustible tobacco (defined as cigars, cigarillos, or little cigars; pipes filled with tobacco; bidis; tobacco in a hookah or waterpipe), smokeless/non-combustible tobacco (chewing tobacco, snuff, or dip; snus; heated tobacco products; nicotine pouches; other oral nicotine products), and e-cigarettes. Use of heated tobacco products was assessed from 2019 and nicotine pouches from 2021. The 2023 survey replaced questions assessing use of dissolvable tobacco products with ones assessing use of oral nicotine products more broadly (defined as lozenges, discs, tablets, gums, dissolvable tobacco products, and other products).

#### Nicotine dependence

Nicotine dependence was assessed using measures of craving and time to wanting to first use tobacco products after waking. These questions were generally asked with guidance making it clear to respondents that ‘tobacco products’ included e-cigarettes (although they do not contain tobacco, e-cigarettes are regulated as a tobacco product in the US^20^). For example, in 2023, the questions were prefaced with the following instructions: ‘In answering the next 5 questions, please think about all of the tobacco products that you have used in the past 30 days, including e-cigarettes, cigarettes, cigars, smokeless tobacco, snus, nicotine pouches, other oral nicotine products, hookahs, heated tobacco products, pipe tobacco, bidis, and roll-your-own cigarettes.’ Nonetheless, we include a sensitivity analysis exploring the potential impact of any underreporting by participants who used e-cigarettes only who may have mistakenly thought these questions did not apply to them.

Craving was assessed with the question: ‘During the past 30 days, have you had a strong craving or felt like you really needed to use a tobacco product of any kind?’ Participants were coded 1 if they responded ‘yes’, else they were coded 0. In 2014, examples were given at the end of the question: ‘such as smoking a cigarette or cigar, or using chewing tobacco’. No examples were given in subsequent surveys.

Time to wanting to first use tobacco products after waking was assessed with the question: ‘How soon after you wake up do you want to use a tobacco product?’ Response options were: (a)I do not want to use tobacco, (b) within 5 minutes, (c) from 6 to 30 minutes, (d) from more than 30 minutes to 1 hour, (e) after more than 1 hour but less than 24 hours, (f) I rarely want to use tobacco. Participants were coded 1 if they reported wanting to first use tobacco products within 30 minutes of waking (responses a and b), else they were coded 0.

Both craving and time to first use after waking are validated and widely employed measures of nicotine dependence among adults.^21^

These questions were asked to all participants up to 2019, but from 2020 they were only asked to those who reported using one or more products in the past 30 days. For consistency across the time series, we imputed these variables as 0 (i.e., not dependent) for non-users in each annual survey. Note this deviates from the approach we took in our previous analysis of data collected up to 2019.^8^

### Statistical analysis

Data were analysed using R v.4.4.1. We used the *survey* package to account for the complex sampling design.

We calculated estimates (with 95% confidence intervals [CIs]) of the prevalence of past-30-day use of different nicotine products by survey year, and of the prevalence of nicotine dependence (indexed by past-30-day craving and wanting to first use tobacco products within 30 minutes of waking) in relation to survey year and past-30-day product use.

To explore the potential impact of underestimation of dependence among participants who reported using e-cigarettes only (on the basis that they may not have understood that the questions applied to them, given e-cigarettes do not contain tobacco^9^), we recalculated estimates of the population burden of nicotine dependence assigning those using only e-cigarettes the same level of dependence as cigarette smokers (the product typically associated with the highest level of dependence^8^). For each symptom of dependence, we recalculated the population burden of nicotine dependence within each survey year as follows:

1. Calculated the proportion of the observed estimate of the population burden of nicotine dependence attributable to those using e-cigarettes only: % *using ecigarettes only* × % *reporting symptom of dependence among those using ecigarettes only*.
2. Re-estimated this proportion assuming e-cigarettes were as dependence-forming as cigarettes: % *using ecigarettes only* × % *reporting symptom of dependence among those using cigarettes*.
3. Calculated the uplift to be applied to the observed estimate of the population burden of nicotine dependence: *result of step 2 – result of step* 1.
4. Applied the uplift to the observed estimate of the population burden of nicotine dependence: *observed estimate of population burden of nicotine dependence + result of step* 3.

We plotted these re-estimated values alongside observed estimates to provide an upper bound for the estimate of the population burden of nicotine dependence (i.e., the total proportion of high-school students, including those who reported not having used any product in the past 30 days, experiencing symptoms of nicotine dependence).

## Results

### Nicotine product use

**Figure 1** shows patterns of past-30-day nicotine product use among high-school students by survey year. Estimates with 95% CIs are provided in **Table S1**.

**Figure 1.**
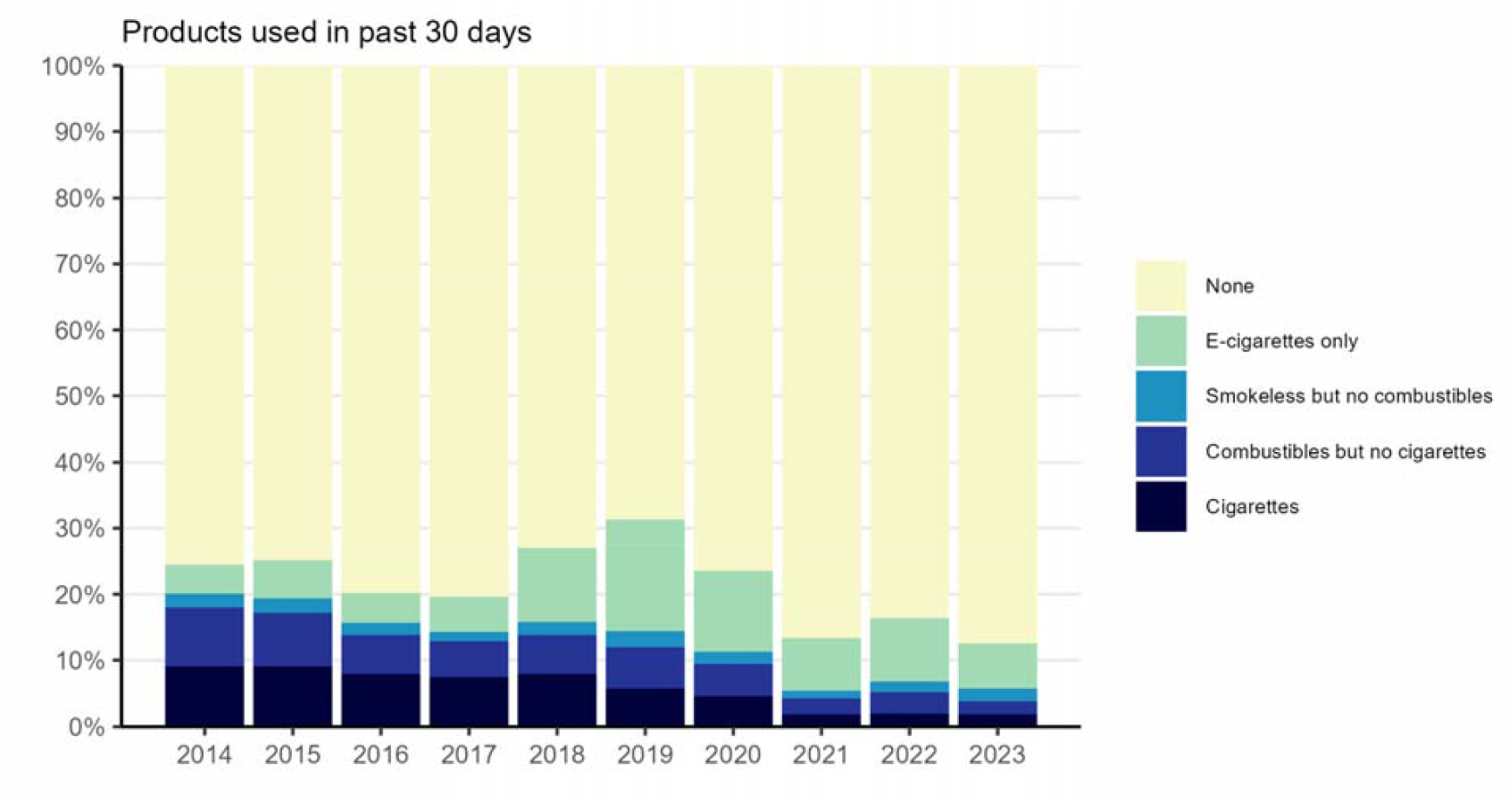
Past-30-day nicotine product use among US high-school students, 2014 to 2023. Unweighted sample sizes: 2014 *n*=11,399; 2015 *n*=9,433; 2016 *n*=10,897; 2017 *n*=10,186; 2018 *n*=10,991; 2019 *n*=10,097; 2020 *n*=7,453; 2021 *n*=10,515; 2022 *n*=16,118; 2023 *n*=10,879. Smokeless includes chewing tobacco, snuff, or dip; snus; heated tobacco products; nicotine pouches; other oral nicotine products. Combustibles includes cigars, cigarillos, or little cigars; pipes filled with tobacco; bidis; tobacco in a hookah or waterpipe. Estimates with 95% CIs are provided in **Table S1**.

The prevalence of nicotine product use changed non-linearly across the period. The proportion of high-school students who reported having used any nicotine product in the past 30 days decreased between 2014 and 2017, from 24.5% [22.5–26.6%] to 19.6% [16.8–22.4%]. There was then a sharp increase in prevalence between 2017 and 2019, reaching a high of 31.4% [29.0–33.7%] in 2019. This trend reversed between 2019 and 2023, with prevalence falling to a new low of 12.5% [10.9–14.1%] by 2023.

There were also changes in the types of products being used. The proportion of high-school students who reported using cigarettes decreased over time, from 9.0% [7.9–10.3%] in 2014 to 1.8% [1.5–2.3%] in 2021 and remained at this level in 2023 (1.8% [1.4–2.4%]). There was a similar decline in the proportion who reported using other combustible tobacco products (but not cigarettes), from 9.0% [8.1–10.0%] in 2014 to 2.5% [1.9–3.1%] in 2021 and 2.0% [1.5–2.6%] in 2023. As a result, the overall proportion using any combustible tobacco products fell from 18.0% [16.6–19.4%] in 2014 to 4.3% [3.5–5.1%] in 2021 and remained relatively stable up to 2023 (3.9% [3.2–4.5%]). The proportion who reported using smokeless/non-combustible products but no combustibles was consistently low (ranging between 1.2% and 2.4%). The proportion who reported using e-cigarettes only was roughly stable between 2014 and 2017 (ranging between 4.4% and 5.8%), then increased rapidly to a high of 17.0% [15.3–18.7%] in 2019, before falling to 6.7% [5.6–7.9%] by 2023.

### Nicotine dependence

**Figure 2** shows the proportion of all high-school students reporting symptoms of nicotine dependence by survey year. **Figure 3** shows levels of dependence among those using different categories of nicotine products by survey year. Estimates with 95% CIs are provided in **Table S2** (past-30-day craving) and **Table S3** (wanting to use within 30 minutes of waking).

**Figure 2.**
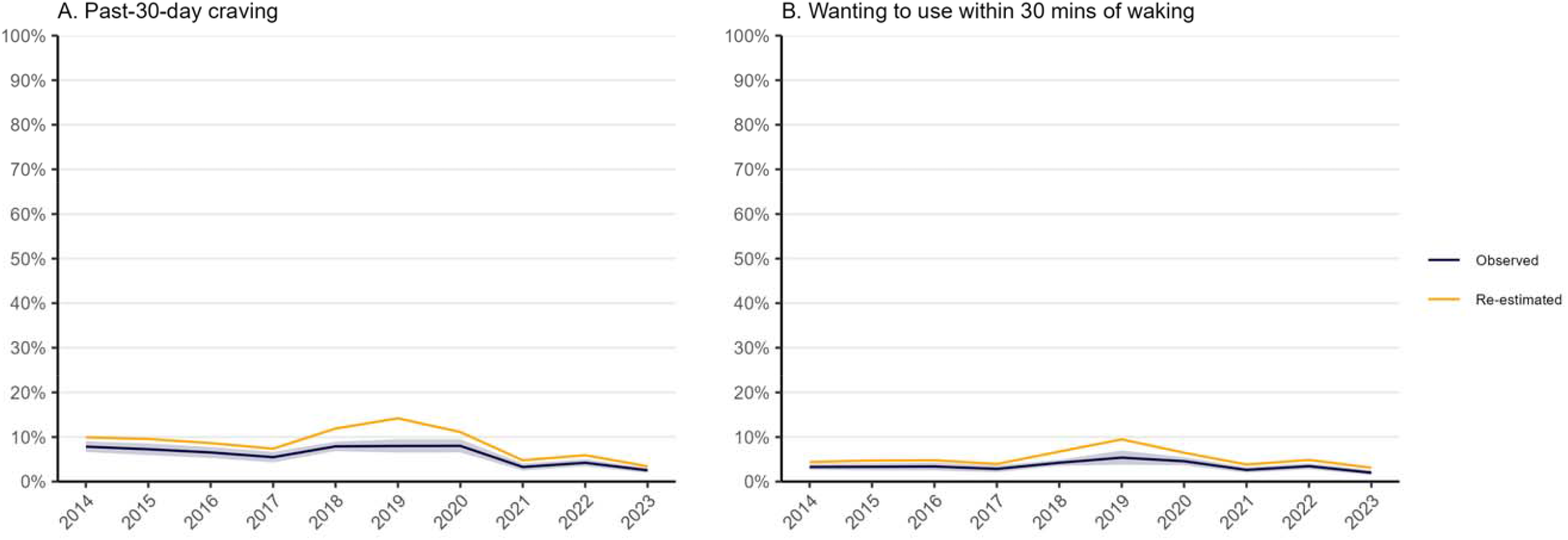
Nicotine dependence among US high-school students, 2014 to 2023. Observed values are estimates of the proportion (with 95% confidence interval) who reported symptoms of nicotine dependence among all participants surveyed (including those who did not report any past-30-day nicotine product use). Re-estimated values account for potential underestimation of nicotine dependence among participants who used e-cigarettes only (on the basis that they may not have thought questions about tobacco products applied to them), assuming that e-cigarettes were as dependence-forming as cigarettes (a plausible maximum level of dependence; see **Table S4** and **Table S5** for details of re-estimation calculations). Estimates assume no dependence among participants who reported no past-30-day nicotine product use. Sample sizes and observed estimates with 95% CIs are provided in **Table S2** (past-30-day craving) and **Table S3** (wanting to use within 30 mins of waking).

**Figure 3.**
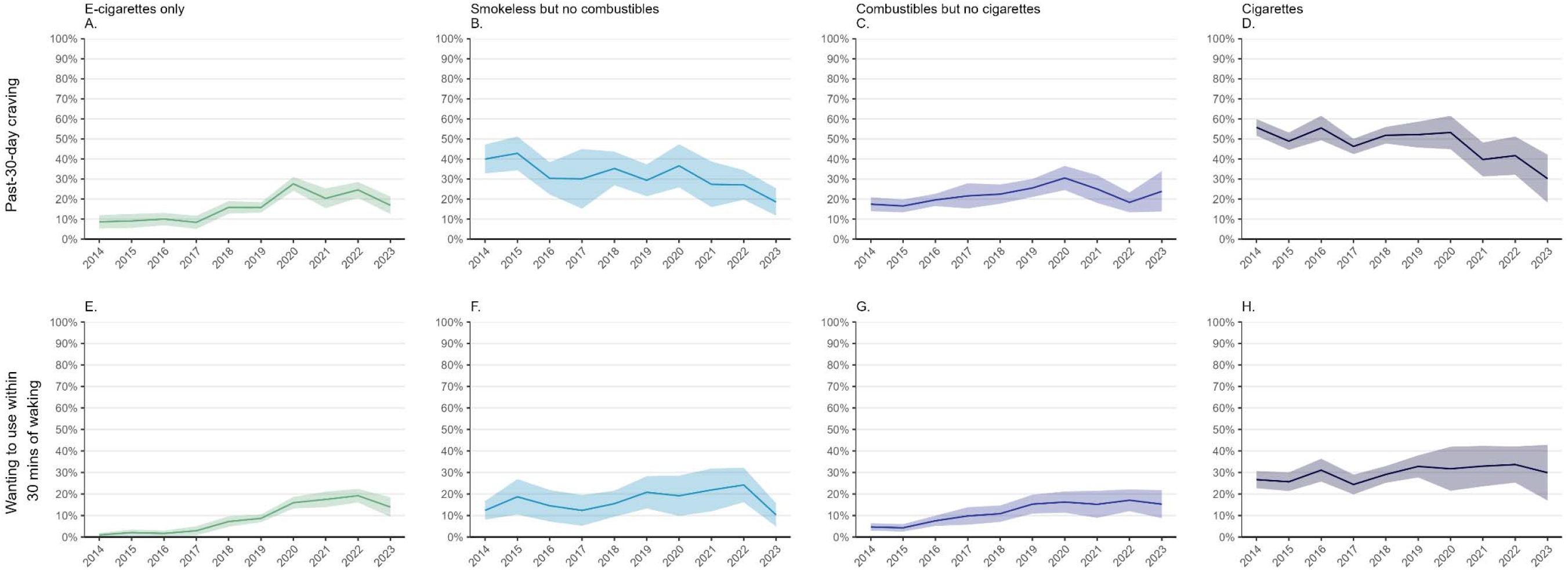
Nicotine dependence among US high-school students by product used, 2014 to 2023. Data shown are estimates of the proportions (with 95% confidence intervals) of participants using different categories of nicotine products who reported symptoms of nicotine dependence. Sample sizes and estimates with 95% CIs are provided in **Table S2** (past-30-day craving) and **Table S3** (wanting to use within 30 mins of waking).

Across the period, the proportion of high-school students reporting symptoms of nicotine dependence was consistently substantially lower than the proportion who reported past-30-day use of any tobacco products. However, changes over time followed a broadly similar pattern. Between 2014 and 2017, the proportion who reported strong craving to use a tobacco product of any kind in the past 30 days decreased from 7.8% [6.6–9.0%] to 5.5% [4.3–6.7%] (note the slight overlap in 95% CIs, indicating some uncertainty). It then increased to 7.9% [6.8–9.0%] between 2017 and 2018 and remained stable up to 2020, before falling sharply to 3.3% [2.5– 4.1%] in 2021 and then to a new low of 2.5% [1.9–3.1%] by 2023 (**Figure 2A**). Results were similar for the proportion who reported wanting to first use a tobacco product within 30 minutes of waking: little change between 2014 (3.3% [2.6–3.9%]) and 2017 (2.8% [2.1–3.6%]), followed by an increase to 5.4% [3.8–7.0%] by 2019 and subsequent decrease to 2.0% [1.4–2.6%] by 2023 (**Figure 2B**).

The proportion of high-school students reporting symptoms of nicotine dependence was consistently highest among those who reported using cigarettes in the past 30 days compared with those who used other combustibles, smokeless/non-combustible products, or e-cigarettes only (**Figure 3**). However, there was a notable increase in symptoms of nicotine dependence among those using e-cigarettes only across the period. The proportion of those who used e-cigarettes only who reported strong craving to use a tobacco product of any kind in the past 30 days was relatively low and stable (ranging between 8.4% and 10.0%) between 2014 and 2017, increased to 27.6% [24.2–31.1%] by 2020, and then decreased to 16.9% [12.5–21.3%] by 2023 (**Figure 3A**). The proportion who reported wanting to use a tobacco product within 30 minutes of waking increased was also consistently low (ranging between 1.0% and 2.9%) between 2014 and 2017, then increased steadily to a high of 19.2% [16.0–22.4%] by 2022; this was followed by an uncertain decrease to 13.9% [9.4–18.5%] in 2023 (**Figure 3E**).

When we re-estimated the total population burden of nicotine dependence to account for potential underreporting of symptoms among those using e-cigarettes only (see **Table S4** and **Table S5** for calculations), changes over time followed a similar pattern but were more pronounced (**Figure 2**). Compared with estimates based on the observed data, the peak in 2019 was almost twice as high, reaching 14.2% for past-30-day craving and 9.5% for wanting to use within 30 minutes of waking, but values in 2023 were only slightly higher at 3.4% and 3.1%, respectively (compared with observed values of 2.5% [1.9–3.1%] and 2.0% [1.4–2.6%]).

## Discussion

In a previous study that analysed NYTS data up to 2019, we concluded that increases in the prevalence of nicotine product use (in particular, e-cigarettes) among US high-school students did not appear to have been accompanied by a similar increase in the population burden of nicotine dependence.^8^ The present analyses, which include four additional years of data, show that the proportion of high-school students using any nicotine product peaked in 2019 and has since fallen steeply to a record low in 2023. Use of cigarettes and other combustible products declined steadily between 2014 and 2021 then was stable up to 2023, while e-cigarette use spiked between 2017 and 2019 before decreasing by 2023. The overall burden of nicotine dependence showed a similar pattern, with a peak in 2019 and a notable reduction by 2023.

Vaping is a contentious issue. Evidence from high-quality randomised controlled trials shows e-cigarettes are effective for helping people to stop smoking.^22,23^ This is broadly (although not universally) accepted by researchers, public health organisations, and policymakers. However, there is widespread concern that e-cigarettes are recruiting people into nicotine dependence who would not otherwise have used any nicotine or tobacco,^5^ and that they have provided a gateway to more harmful cigarette smoking.^24^ This has led many countries to impose strict regulations on vaping products,^25^ which has probably contributed to reduced uptake among young people but potentially undermined their usefulness for smoking cessation. Our results show no evidence of a gateway effect from vaping to smoking at the population level: despite a sharp rise in e-cigarette use in 2019, use of cigarettes and other combustible tobacco products continued to fall and in 2023 were lower than ever. Likewise, the spike in e-cigarette use among high-school students did not lead to a sustained increase in the population burden of nicotine dependence, which is now at a historically low level.

We found that while dependence symptoms were consistently highest among cigarette users, they were increasingly prevalent among those using e-cigarettes only after 2017. This is consistent with data from other surveys^9^ and experimental studies^17,18^ showing e-cigarette devices have become more efficient at delivering nicotine over time – particularly since the introduction of nicotine salts.^26^ In our analyses, we explored the possibility that some e-cigarette users may not have interpreted nicotine dependence questions as applicable to them (even though guidance clarified this). If this were true, it could have led us to underestimate dependence among those using only e-cigarettes. However, even when we assumed e-cigarettes were as dependence-forming as cigarettes, our results did not show a sustained increase in the population burden of dependence. This evidence directly contradicts claims that e-cigarettes are creating a new generation of people addicted to nicotine.^5^

Collectively, our findings show that rises in youth vaping may not necessarily be sustained in the long-term; rates declined sharply following the peak in 2019, much more rapidly than typically seen for smoking. This might mean youth vaping is less difficult to address than smoking, assuming recent product developments (e.g., additions of synthetic coolants) do not make e-cigarettes more dependence-forming and difficult to quit. Therefore, a proportionate approach to regulation might aim to reduce the access and appeal of e-cigarettes to young people (e.g., targeting restrictions at marketing and branding), without jeopardising their effectiveness for smoking cessation (e.g., limiting nicotine content). Careful monitoring of trends in youth use and dependence can allow restrictions to be adapted if and when needed.

Finally, we note that prevalence of use of cigarettes and other combustible tobacco products appears to have stalled since 2021 following years of steady decline. Additional policies may be required to address the final ∼4% of high-school students smoking combustible tobacco.

This study had several limitations. First, nicotine product use was determined based on self-reports of any use within the past 30 days. Future studies could offer further insights into changing patterns of dependence by accounting for frequency of use. Secondly, for surveys conducted from 2020 onward, nicotine dependence questions were only asked to participants who had used a nicotine product in the past 30 days. We coded dependence as 0 (i.e., not dependent) for non-users, which might underestimate dependence for individuals who use nicotine products infrequently. Thirdly, the survey did not differentiate between nicotine-containing and nicotine-free e-cigarettes, or different types of devices (e.g., older vs. newer salt-based nicotine e-cigarettes), which may have varying impacts on dependence. Finally, the repeat cross-sectional design cannot assess causal relationships between changes in (or continued) product use and nicotine dependence over time. Longitudinal data are needed to capture individual trajectories of dependence.

In conclusion, rapid increases in the prevalence of nicotine product use (in particular, e-cigarettes) among US high-school students in the late 2010s were short-lived and were not accompanied by a sustained increase in the population burden of nicotine dependence. By 2023, both nicotine product use and nicotine dependence were at historic low levels.

## Data Availability

Data are available online at https://www.cdc.gov/tobacco/about-data/surveys/national-youth-tobacco-survey.html

## References

1 Centers for Disease Control and Prevention. Youth and Tobacco Use. Cent. Dis. Control Prev. 2019.https://www.cdc.gov/tobacco/data_statistics/fact_sheets/youth_data/tobacco_use/index.htm (accessed 18 Dec2019).

2 Mejia MC, Adele A, Levine RS, Hennekens CH, Kitsantas P. Trends in Cigarette Smoking Among United States Adolescents. Ochsner J 2023; 23: 289–295.

3 Nelson DE, Mowery P, Asman K, Pederson LL, O’Malley PM, Malarcher A et al. Long-Term Trends in Adolescent and Young Adult Smoking in the United States: Metapatterns and Implications. Am J Public Health 2008; 98: 905–915.

4 Sun R, Mendez D, Warner KE. Trends in Nicotine Product Use Among US Adolescents, 1999-2020. JAMA Netw Open 2021; 4: e2118788.

5 Matthews J, Matthews M, Cherian V. A cloud of addiction: how vaping has created a new generation of addicts. Br J Gen Pract 2023; 73. doi:10.3399/bjgp23X734325.

6 Wang TW, Gentzke AS, Creamer MR, Cullen KA, Holder-Hayes E, Sawdey MD et al. Tobacco Product Use and Associated Factors Among Middle and High School Students - DUnited States, 2019. Morb Mortal Wkly Rep Surveill Summ Wash DC 2002 2019; 68: 1–22.

7 Wang TW, Gentzke A, Sharapova S, Cullen KA, Ambrose BK, Jamal A. Tobacco Product Use Among Middle and High School Students - United States, 2011–2017. MMWR Morb Mortal Wkly Rep 2018; 67: 629–633.

8 Jackson SE, Brown J, Jarvis MJ. Dependence on nicotine in US high school students in the context of changing patterns of tobacco product use. Addiction 2021; 116: 1859–1870.

9 Gomes MN, Reid JL, Rynard VL, East KA, Goniewicz ML, Piper ME et al. Comparison of Indicators of Dependence for Vaping and Smoking: Trends Between 2017 and 2022 Among Youth in Canada, England, and the United States. Nicotine Tob Res 2024; : tae060.

10 Centers for Disease Control and Prevention. Outbreak of Lung Injury Associated with the Use of E-Cigarette, or Vaping, Products. 2020.https://www.cdc.gov/tobacco/basic_information/e-cigarettes/severe-lung-disease.html#latest-outbreak-information (accessed 28 Jun2023).

11 Blount BC, Karwowski MP, Shields PG, Morel-Espinosa M, Valentin-Blasini L, Gardner M et al. Vitamin E Acetate in Bronchoalveolar-Lavage Fluid Associated with EVALI. N Engl J Med 2020; 382: 697–705.

12 East K, Reid JL, Burkhalter R, Wackowski OA, Thrasher JF, Tattan-Birch H et al. Exposure to Negative News Stories About Vaping, and Harm Perceptions of Vaping, Among Youth in England, Canada, and the United States Before and After the Outbreak of E-cigarette or Vaping-Associated Lung Injury (‘EVALI’). Nicotine Tob Res 2022; 24: 1386–1395.

13 Siegel M, Katchmar A. Effect of flavored e-cigarette bans in the United States: What does the evidence show? Prev Med 2022; 165: 107063.

14 Koh HK, Douglas CE. The San Francisco Ban and the Future of e-Cigarettes. JAMA 2019; 322: 1540–1541.

15 Jun J, Kim JK. Do state regulations on e-cigarettes have impacts on the e-cigarette prevalence? Tob Control 2021; 30: 221–226.

16 Birdsey J, Cornelius M, Jamal A, Park-Lee E, Cooper MR, Wang J et al. Tobacco Product Use Among U.S. Middle and High School Students — National Youth Tobacco Survey, 2023. MMWR Morb Mortal Wkly Rep 2023; 72. doi:10.15585/mmwr.mm7244a1.

17 Talih S, Salman R, Soule E, El-Hage R, Karam E, Karaoghlanian N et al. Electrical features, liquid composition and toxicant emissions from ‘pod-mod’-like disposable electronic cigarettes. Tob Control 2022; 31: 667–670.

18 Leventhal AM, Madden DR, Peraza N, Schiff SJ, Lebovitz L, Whitted L et al. Effect of Exposure to e-Cigarettes With Salt vs Free-Base Nicotine on the Appeal and Sensory Experience of Vaping: A Randomized Clinical Trial. JAMA Netw Open 2021; 4: e2032757.

19 Office on Smoking and Health. 2023 National Youth Tobacco Survey: Methodology Report. US Department of Health and Human Services, Centers for Disease Control and Prevention, National Center for Chronic Disease Prevention and Health Promotion, Office on Smoking and Health: Atlanta, GA, 2024chrome-extension://efaidnbmnnnibpcajpcglclefindmkaj/ https://www.cdc.gov/tobacco/data_statistics/surv eys/nyts/pdfs/2023_NYTS_Methodology_508.pdf (accessed 8 Aug2024).

20 US Food and Drug Administration (FDA). Tobacco Products. FDA. 2024.https://www.fda.gov/tobacco-products (accessed 8 Aug2024).

21 Fagerström K. Time to first cigarette; the best single indicator of tobacco dependence? Monaldi Arch Chest Dis Arch Monaldi Mal Torace 2003; 59: 91.

22 Lindson N, Butler AR, McRobbie H, Bullen C, Hajek P, Begh R et al. Electronic cigarettes for smoking cessation. Cochrane Database Syst Rev 2024. doi:10.1002/14651858.CD010216.pub8.

23 Auer R, Schoeni A, Humair J-P, Jacot-Sadowski I, Berlin I, Stuber MJ et al. Electronic Nicotine-Delivery Systems for Smoking Cessation. N Engl J Med 2024; 390: 601–610.

24 Shahab L, Brown J, Boelen L, Beard E, West R, Munafò MR. Unpacking the Gateway Hypothesis of E-Cigarette Use: The Need for Triangulation of Individual- and Population-Level Data. Nicotine Tob Res 2022; 24: 1315–1318.

25 Warner KE, Benowitz NL, McNeill A, Rigotti NA. Nicotine e-cigarettes as a tool for smoking cessation. Nat Med 2023; 29: 520–524.

26 Cho YJ, Mehta T, Hinton A, Sloan R, Nshimiyimana J, Tackett AP et al. E-Cigarette Nicotine Delivery Among Young Adults by Nicotine Form, Concentration, and Flavor: A Crossover Randomized Clinical Trial. JAMA Netw Open 2024; 7: e2426702.

27 Buss V, Kock L, West R, Kale D, Brown J. Trends in electronic cigarette use in England. 2024.https://smokinginengland.info/graphs/e-cigarettes-latest-trends.

28 Lindson N, Theodoulou A, Ordóñez-Mena JM, Fanshawe TR, Sutton AJ, Livingstone-Banks J et al. Pharmacological and electronic cigarette interventions for smoking cessation in adults: component network meta-analyses. Cochrane Database Syst Rev 2023. doi:10.1002/14651858.CD015226.pub2.

