## Supplementary material for "Changing patterns of nicotine product use and nicotine dependence among US high school students: the National Youth Tobacco Survey, 2014-2023": Table S1

**Table S1.** Past-30-day nicotine product use among US high school students, 2014 to 2023

| | | % [95% CI] reporting past-30-day use of… | | | | |
|---|---|---|---|---|---|---|
| **Year** | ***N*** | **E-cigarettes only** | **Smokeless but no combustibles**[1] | **Combustibles but no cigarettes**[2] | **Cigarettes** | **Any nicotine product** |
| 2014 | 11,399 | 4.4 [3.4–5.6] | 2.1 [1.6–2.7] | 9.0 [8.1–10.0] | 9.0 [7.9–10.3] | 24.5 [22.5–26.6] |
| 2015 | 9,433 | 5.8 [4.9–7.0] | 2.2 [1.6–3.1] | 8.1 [7.2–9.2] | 9.1 [7.7–10.7] | 25.2 [23.1–27.4] |
| 2016 | 10,897 | 4.6 [3.8–5.5] | 1.8 [1.4–2.4] | 6.0 [5.2–6.8] | 7.8 [6.5–9.4] | 20.2 [18.2–22.2] |
| 2017 | 10,186 | 5.4 [4.2–6.8] | 1.4 [1.0–2.0] | 5.4 [4.6–6.4] | 7.4 [6.1–9.0] | 19.6 [16.8–22.4] |
| 2018 | 10,991 | 11.2 [9.6–13.1] | 1.9 [1.5–2.5] | 5.9 [5.1–6.9] | 7.9 [6.8–9.2] | 27.0 [25.0–28.9] |
| 2019 | 10,097 | 17.0 [15.3–18.7] | 2.4 [1.8–3.2] | 6.2 [5.2–7.3] | 5.8 [4.5–7.4] | 31.4 [29.0–33.7] |
| 2020 | 7,453 | 12.3 [10.9–14.0] | 1.9 [1.4–2.5] | 4.8 [4.1–5.7] | 4.6 [3.5–6.0] | 23.6 [21.0–26.3] |
| 2021 | 10,515 | 7.9 [6.6–9.3] | 1.2 [0.9–1.6] | 2.5 [1.9–3.1] | 1.8 [1.5–2.3] | 13.3 [11.4–15.2] |
| 2022 | 16,118 | 9.7 [8.6–10.9] | 1.5 [1.2–2.0] | 3.2 [2.6–3.9] | 2.0 [1.6–2.5] | 16.4 [14.6–18.3] |
| 2023 | 10,879 | 6.7 [5.6–7.9] | 2.0 [1.5–2.5] | 2.0 [1.5–2.6] | 1.8 [1.4–2.4] | 12.5 [10.9–14.1] |

[1] Includes chewing tobacco, snuff, or dip; snus; heated tobacco products; nicotine pouches; other oral nicotine products.

[2] Includes cigars, cigarillos, or little cigars; pipes filled with tobacco; bidis; tobacco in a hookah or waterpipe.

**Table S2.** Past-30-day craving to use tobacco products among US high school students, overall and by product use, 2014 to 2023

| | | % [95% CI] reporting strong craving to use a tobacco product in the past 30 days among those using… | | | | |
|---|---|---|---|---|---|---|
| **Year** | ***N*** | **E-cigarettes only** | **Smokeless but no combustibles[1]** | **Combustibles but no cigarettes[2]** | **Cigarettes** | **Total population[3]** |
| 2014 | 11,399 | 8.7 [5.3–12.0] | 40.0 [32.8–47.2] | 17.4 [14.0–20.9] | 55.8 [51.6–59.9] | 7.8 [6.6–9.0] |
| 2015 | 9,433 | 9.1 [5.5–12.6] | 42.8 [34.4–51.3] | 16.6 [13.4–19.8] | 48.9 [44.5–53.2] | 7.2 [5.9–8.6] |
| 2016 | 10,897 | 10.0 [6.9–13.1] | 30.3 [22.3–38.4] | 19.6 [16.5–22.6] | 55.5 [49.4–61.6] | 6.5 [5.3–7.8] |
| 2017 | 10,186 | 8.4 [5.1–11.7] | 30.1 [15.2–45.0] | 21.6 [15.3–27.9] | 46.3 [42.4–50.1] | 5.5 [4.3–6.7] |
| 2018 | 10,991 | 15.9 [12.7–19.0] | 35.3 [26.9–43.7] | 22.5 [17.7–27.2] | 51.8 [47.7–56.0] | 7.9 [6.8–9.0] |
| 2019 | 10,097 | 15.8 [13.2–18.4] | 29.4 [21.4–37.3] | 25.5 [21.0–30.0] | 52.2 [45.7–58.6] | 8.0 [6.5–9.5] |
| 2020 | 7,453 | 27.6 [24.2–31.1] | 36.6 [25.8–47.4] | 30.5 [24.5–36.6] | 53.2 [44.9–61.5] | 8.0 [6.5–9.5] |
| 2021 | 10,515 | 20.3 [15.4–25.3] | 27.4 [16.0–38.8] | 25.0 [18.1–31.9] | 39.7 [31.3–48.2] | 3.3 [2.5–4.1] |
| 2022 | 16,118 | 24.6 [20.6–28.5] | 27.1 [19.7–34.4] | 18.3 [13.4–23.3] | 41.7 [32.1–51.2] | 4.2 [3.4–5.0] |
| 2023 | 10,879 | 16.9 [12.5–21.3] | 18.5 [11.7–25.4] | 23.9 [13.8–33.9] | 30.2 [18.2–42.2] | 2.5 [1.9–3.1] |

[1] Includes chewing tobacco, snuff, or dip; snus; heated tobacco products; nicotine pouches; other oral nicotine products.
[2] Includes cigars, cigarillos, or little cigars; pipes filled with tobacco; bidis; tobacco in a hookah or waterpipe.
[3] Includes participants reporting no past-30-day use of any nicotine product.

**Table S3.** Wanting to use a tobacco product within 30 minutes of waking among US high school students, overall and by product use, 2014 to 2023

| | | % [95% CI] reporting wanting to use a tobacco product within 30 minutes of waking among those using… | | | | |
|---|---|---|---|---|---|---|
| **Year** | ***N*** | **E-cigarettes only** | **Smokeless but no combustibles[1]** | **Combustibles but no cigarettes[2]** | **Cigarettes** | **Total population[3]** |
| 2014 | 11,399 | 1.0 [0.0–1.9] | 12.4 [8.0–16.7] | 4.7 [2.9–6.4] | 26.7 [22.7–30.7] | 3.3 [2.6–3.9] |
| 2015 | 9,433 | 2.0 [0.6–3.5] | 18.7 [10.5–26.9] | 4.3 [2.5–6.0] | 25.7 [21.4–30.0] | 3.3 [2.5–4.1] |
| 2016 | 10,897 | 1.7 [0.4–3.0] | 14.5 [7.2–21.9] | 7.5 [5.2–9.9] | 31.1 [25.8–36.4] | 3.4 [2.5–4.3] |
| 2017 | 10,186 | 2.9 [0.8–5.0] | 12.4 [5.2–19.5] | 9.8 [5.7–13.9] | 24.4 [19.8–29.0] | 2.8 [2.1–3.6] |
| 2018 | 10,991 | 7.2 [4.8–9.6] | 15.5 [9.4–21.6] | 10.8 [7.0–14.7] | 29.1 [25.2–33.0] | 4.2 [3.5–4.9] |
| 2019 | 10,097 | 8.7 [6.9–10.5] | 20.8 [13.3–28.3] | 15.3 [10.9–19.7] | 32.8 [27.7–38.0] | 5.4 [3.8–7.0] |
| 2020 | 7,453 | 15.9 [13.2–18.6] | 19.2 [9.8–28.6] | 16.2 [11.3–21.2] | 31.7 [21.5–42.0] | 4.6 [3.7–5.5] |
| 2021 | 10,515 | 17.5 [13.8–21.1] | 21.9 [11.9–31.8] | 15.2 [8.9–21.5] | 33.0 [23.6–42.4] | 2.6 [2.0–3.2] |
| 2022 | 16,118 | 19.2 [16.0–22.4] | 24.2 [16.1–32.2] | 17.1 [12.0–22.1] | 33.7 [25.3–42.1] | 3.5 [2.8–4.1] |
| 2023 | 10,879 | 13.9 [9.4–18.5] | 10.3 [4.8–15.7] | 15.3 [8.7–21.8] | 29.9 [16.9–42.9] | 2.0 [1.4–2.6] |

[1] Includes chewing tobacco, snuff, or dip; snus; heated tobacco products; nicotine pouches; other oral nicotine products.
[2] Includes cigars, cigarillos, or little cigars; pipes filled with tobacco; bidis; tobacco in a hookah or waterpipe.
[3] Includes participants reporting no past-30-day use of any nicotine product.

**Table S4**. Re-estimating the population burden of nicotine dependence assuming e-cigarettes are as dependence-forming as cigarettes: past-30-day craving

| **Year** | **% using e-cigarettes only** | **% reporting past-30-day craving among those using e-cigarettes only** | **% reporting past-30-day craving among those using cigarettes** | **% of population reporting past-30-day craving attributable to e-cigarettes only[1]** | **% of population reporting past-30-day craving attributable to e-cigarettes only if they were as dependence-forming as cigarettes[2]** | **Uplift applied to observed estimate of population burden of dependence[3]** |
|---|---|---|---|---|---|---|
| 2014 | 4.4% | 8.7% | 55.8% | 0.4% | 2.5% | 2.1% |
| 2015 | 5.8% | 9.1% | 48.9% | 0.5% | 2.8% | 2.3% |
| 2016 | 4.6% | 10.0% | 55.5% | 0.5% | 2.6% | 2.1% |
| 2017 | 5.1% | 8.4% | 46.3% | 0.4% | 2.4% | 1.9% |
| 2018 | 11.2% | 15.9% | 51.8% | 1.8% | 5.8% | 4.0% |
| 2019 | 17.0% | 15.8% | 52.2% | 2.7% | 8.9% | 6.2% |
| 2020 | 12.3% | 27.6% | 53.2% | 3.4% | 6.5% | 3.1% |
| 2021 | 7.9% | 20.3% | 39.7% | 1.6% | 3.1% | 1.5% |
| 2022 | 9.7% | 24.6% | 41.7% | 2.4% | 4.0% | 1.7% |
| 2023 | 6.7% | 16.9% | 30.2% | 1.1% | 2.0% | 0.9% |

[1] Estimate based on observed data: % using e-cigarettes only multiplied by % reporting past-30-day craving to use a tobacco product among those using e-cigarettes only.

[2] Re-estimate based on the assumption that e-cigarettes are as dependence-forming as cigarettes: % using e-cigarettes only multiplied by % reporting past-30-day craving to use a tobacco product among those using cigarettes.

[3] Amount the estimate of the population burden of nicotine dependence (based on past-30-day craving) needs to be increased by to account for the re-estimation of dependence among those using e-cigarettes only: % of population reporting past-30-day craving attributable to e-cigarettes only if they were as dependence-forming as cigarettes minus % of population reporting past-30-day craving attributable to e-cigarettes only.

**Table S5**. Re-estimating the population burden of nicotine dependence assuming e-cigarettes are as dependence-forming as cigarettes: wanting to use within 30 minutes of waking

| Year | % using e-cigarettes only | % reporting wanting to use within 30 mins among those using e-cigarettes only | % reporting wanting to use within 30 mins among those using cigarettes | % of population reporting wanting to use within 30 mins attributable to e-cigarettes only[1] | % of population reporting wanting to use within 30 mins attributable to e-cigarettes only if they were as dependence-forming as cigarettes[2] | Uplift applied to observed estimate of population burden of dependence[3] |
|---|---|---|---|---|---|---|
| 2014 | 4.4% | 1.0% | 26.7% | 0.0% | 1.2% | 1.1% |
| 2015 | 5.8% | 2.0% | 25.7% | 0.1% | 1.5% | 1.4% |
| 2016 | 4.6% | 1.7% | 31.1% | 0.1% | 1.4% | 1.4% |
| 2017 | 5.1% | 2.9% | 24.4% | 0.1% | 1.2% | 1.1% |
| 2018 | 11.2% | 7.2% | 29.1% | 0.8% | 3.3% | 2.5% |
| 2019 | 17.0% | 8.7% | 32.8% | 1.5% | 5.6% | 4.1% |
| 2020 | 12.3% | 15.9% | 31.7% | 2.0% | 3.9% | 1.9% |
| 2021 | 7.9% | 17.5% | 33.0% | 1.4% | 2.6% | 1.2% |
| 2022 | 9.7% | 19.2% | 33.7% | 1.9% | 3.3% | 1.4% |
| 2023 | 6.7% | 13.9% | 29.9% | 0.9% | 2.0% | 1.1% |

[1] Estimate based on observed data: % using e-cigarettes only multiplied by % reporting wanting to use a tobacco product within 30 minutes of waking among those using e-cigarettes only.

[2] Re-estimate based on the assumption that e-cigarettes are as dependence-forming as cigarettes: % using e-cigarettes only multiplied by % reporting wanting to use a tobacco product within 30 minutes of waking among those using cigarettes.

[3] Amount the estimate of the population burden of nicotine dependence (based on wanting to use within 30 minutes of waking) needs to be increased by to account for the re-estimation of dependence among those using e-cigarettes only: % of population reporting wanting to use within 30 minutes attributable to e-cigarettes only if they were as dependence-forming as cigarettes minus % of population reporting wanting to use within 30 minutes attributable to e-cigarettes only.
